# Quality of Human Expert vs. Large Language Model Generated Multiple Choice Questions in the Field of Mechanical Ventilation

**DOI:** 10.1101/2025.03.16.25324082

**Authors:** Sami Safadi, Roxana Amirahmadi, Abdulhakim Tlimat, Randal Rovinski, Junfeng Sun, Burton W. Lee, Nitin Seam, the Critical Care Education Research Consortium

## Abstract

**Background:** Mechanical ventilation (MV) is a critical competency in critical care training, yet standardized methods for assessing MV-related knowledge are lacking. Traditional multiple-choice question (MCQ) development is resource-intensive, and prior studies have suggested that generative AI tools could streamline question creation. However, the effectiveness and reliability of AI-generated MCQs remain unclear. This study evaluates whether MCQs generated by ChatGPT are non-inferior to human-expert (HE) created questions in terms of quality and relevance for MV education.

**Methods:** Three key MV topics were selected: Equation of Motion & Ohm’s Law, Tau & Auto PEEP, and Oxygenation. Fifteen learning objectives were used to generate 15 AI-written MCQs via a standardized prompt with ChatGPT o1 (model o1-preview-2024-09-12). A group of 31 faculty experts, all of whom regularly teach MV, evaluated both AI-generated and HE-generated MCQs. Each MCQ was assessed based on its alignment with learning objectives, accuracy, clarity, plausibility of distractors, and difficulty level. The faculty members were blinded to the provenance of the MCQ questions. The non-inferiority margin was predefined as 15% of the total possible score (−3.45).

**Results:** AI-generated MCQs were statistically non-inferior to expert-written MCQs (95% upper CI: [-1.15, ∞]). Additionally, respondents were unable to reliably differentiate AI-generated from HE-written MCQs (p = 0.32).

**Conclusion:** AI-generated MCQs using ChatGPT o1 are comparable in quality and difficulty to those written by human experts. Given the time and resource-intensive nature of human MCQ development, AI-assisted question generation may serve as an efficient and scalable alternative for medical education assessment, even in highly specialized domains such as mechanical ventilation.

## Background and Objectives

Safe and effective use of mechanical ventilation (MV) requires the clinician to master a wide range of complex topics in respiratory physiology and disease mechanisms. Traditional methods of evaluating knowledge have relied heavily on using multiple-choice questions (MCQs).

Furthermore, while management of MV is a core competency in critical care training [1], standard and effective methods for assessment of these highly technical skills are lacking. A key challenge in the development of effective assessments is that the process of question generation can be time-consuming and resource-intensive for educators. Generative AI tools offer an opportunity to streamline and enhance this essential task for educators. This study aims to investigate the viability and efficacy of employing a large language model (LLM) (specifically ChatGPT model “o1-preview-2024-09-12”) to generate high-quality MCQs compared to the “gold standard” questions created by human expert educators in mechanical ventilation.

LLMs like GPT-4 and Med-PaLM have demonstrated significant potential and versatility across various fields, particularly in natural language processing tasks such as summarization, translation, code synthesis, and logical reasoning. This technological prowess has sparked interest in their application within the medical domain. [2] Some applications include case diagnoses, medical examinations, and assessing consistency with medical guidelines. [3-6] Despite its promise, the integration of LLMs into clinical practice has limitations, notably in diagnosing complex cases and maintaining consistency with established medical guidelines. The variable and sometimes inaccurate outputs underscore the need for further refinement to enhance their reliability and efficacy in healthcare settings. [7]

The existing body of literature examining the effectiveness of LLMs in generating MCQs remains limited, with mixed findings. In a study conducted by Cheung et al. (2023), 50 MCQs generated by ChatGPT Plus were compared to those created by human educators. [8] Blind assessors evaluated the questions using a standardized scoring rubric. The findings indicated that, while the MCQs produced by ChatGPT Plus were well-structured, their accuracy and depth in assessing knowledge were inconsistent. Furthermore, the relevance of AI-generated questions was rated as the weakest domain compared to human-authored questions.

Unsurprisingly, AI-generated questions were produced significantly faster (20 minutes and 25 seconds) compared to those crafted by humans (211 minutes and 33 seconds). Similarly, Ayub et al. (2023) reported that only 40% of the MCQs generated by ChatGPT version 3.5 were deemed both accurate and appropriate. [9] The variability in the findings across studies may be attributed to methodological differences, such as the version of ChatGPT employed and the assessment criteria used to evaluate the questions. Agarwal et al. (2023) proposed a framework that leveraged deep learning and linguistic techniques to generate MCQs from textual content, with a strong emphasis on ensuring grammatical and semantic accuracy. [10]

ChatGPT o1 is designed to “think” step by step using a technique called “chain of thought reasoning”,[11] which can be thought of as a built-in prompt engineering method. Unlike previous ChatGPT models that typically provide instant answers with minimal internal reasoning, this approach helps the model break down complex problems in a way that resembles human problem-solving. Performance tests have shown that o1 Preview significantly outperforms earlier models, especially in fields requiring strong logical reasoning, such as mathematics, biology, and chemistry. The o1-Preview model’s improved reasoning abilities make it especially well-equipped for complex healthcare tasks, particularly those that demand sophisticated logical analysis and data interpretation.[12]

The main goal of this study is to assess whether AI-generated MCQs are non-inferior to MCQs written by human experts in the field of mechanical ventilation. A secondary goal is to compare AI-generated and human-expert-written MCQs across key criteria: (alignment with learning objectives, accuracy of the correct answer, clarity of the question stem, plausibility of distractor options, and appropriateness of difficulty level).

## Methods

### Writing MCQs

The authors identified three fundamental topics that experts consider essential for teaching MV: Equation of Motion & Ohm’s Law, Tau & Auto PEEP, and Oxygenation. Additionally, they align with the curricular material used by the same authors in their mechanical ventilation training course. For each of the three topics, five learning objectives were identified, totaling 15 objectives (Table 1). These objectives were incorporated into a prompt template and provided to the LLM to generate MCQs.

**Table 1.** Topics and Learning Objectives.

| Topic | Learning Objectives |
| --- | --- |
| **Equation of Motion and Ohm's Law** | • Use a clinical vignette to calculate static compliance<br>• Use a clinical vignette to apply the equation of motion to a patient with high airway pressures<br>• Use a clinical vignette to identify changes in lung static compliance and alveolar pressure<br>• Use a clinical vignette to demonstrate how the equation of motion explains the relationships among alveolar pressure, peak pressure, and P<sub>muscle</sub><br>• Understand the components of the equation of motion in a square wave volume control setting |
| **Tau and Auto PEEP** | • Understand the factors that determine how much volume remains in the lung at the end of expiration<br>• Use a clinical vignette to identify the relationship between compliance, resistance, and risk of auto-PEEP<br>• Recognize the most effective change a clinician can make to acutely treat autoPEEP<br>• Understand that the expiratory time constant represents the time needed to exhale until 37% of tidal volume remains in the lungs<br>• Understand that a patient needs at least 3 taus in expiratory to fully exhale tidal volume |
| **Oxygenation** | • Understand how to calculate the stress index in a mechanically ventilated patient.<br>• Understand oxygen toxicity in a mechanically ventilated patient.<br>• Understand the impact of higher Positive End-Expiratory Pressure (PEEP) on oxygenation and survival in patients with ARDS.<br>• Understand the appropriate application of stress index monitoring for adjusting PEEP in a patient with severe ARDS. Present a clinical vignette<br>• Identify a clinical strategy to improve oxygenation and reduce mortality in ARDS. |

The prompt template used to guide the LLM in creating MCQs was developed and agreed upon by the study authors. The template included the specific objective and instructed the LLM to generate each question with the following components: a question stem, five answer choices, the correct answer, and a brief explanation. Additionally, the prompt specified that the target audience was physicians at the fellow (advanced trainee) level in critical care medicine.

A Python script was used to interact with the ChatGPT model o1-preview-2024-09-12 via the OpenAI API. Each generated multiple-choice question (MCQ) was assigned a unique identifier to track its origin.

Three co-authors, SS, RA, and NS, who are human experts (HE) in mechanical ventilation created 15 multiple-choice questions (MCQs). All three teach MV regionally or nationally as part of their professional responsibilities. To serve as a comparison group, they were given the same objectives given to AI for developing the MCQs. Each expert independently wrote five questions, which were reviewed and approved by the other HE. The AI MCQs were produced after the experts completed their MCQs.

### Survey Design

Survey development followed best practices in educational survey design[13] The survey underwent an iterative review process by the co-authors, who evaluated the structure, response options, and content until consensus was reached. Pre-testing was then conducted using a “think-aloud” approach with a cohort representative of the intended survey population. Based on feedback from pre-testing, the survey was refined iteratively until saturation was achieved. The finalized survey was disseminated via email, with a follow-up reminder email sent over a four-week period, and responses were collected using Google Forms.

The survey evaluated the 15 AI-generated MCQs and 15 human-expert-generated MCQs. The MCQs were presented in random order, and respondents were blinded to the source of each MCQ (i.e., whether it was human-generated or AI-generated). Survey questions were constructed using a 5-point Likert scale, and MCQ quality was based on expert recommendations for best practices for educational MCQ generation. [14,15]:

1. The question aligns with the stated learning objective.
2. The correct answer is indisputably accurate.
3. The question stem is clearly written.
4. The incorrect answer choices are plausible.
5. The question’s difficulty level is appropriate for a graduating critical care fellow.

Respondents rated the first four criteria using a five-point scale from “Strongly Disagree” to “Strongly Agree” and the level-of-difficulty criterion using a separate scale ranging from “Very Easy” to “Very Difficult”. They were also asked to determine whether the MCQ was generated by an HE or AI. To quantify MCQ quality, the first four criteria were scored from 1 to 5, while the difficulty criterion assigned “Appropriate” a score of 3, “Easy” and “Difficult” a score of 2, and “Very Easy” and “Very Difficult” a score of 1, with a maximum possible score of 23 for each MCQ.

### Non-Inferiority Margin

The non-inferiority margin of difference between the scores of AI-generated and expert-generated MCQs was determined through a modified Delphi process. Four co-authors of this study (SS, RA, BWL, and NS) established non-inferiority margins using this process. The non-inferiority margin for the acceptable difference between the AI-generated and HE-generated multiple-choice questions was set to be 15% of the maximum final score, −3.45.

### Statistical Analysis

For the sample size calculation, we assumed the final score to have a common standard deviation of 5, and the expected difference between AI-generated MCQs and expert-generated MCQs to be −2. Using 0.05-level one-sided t test, we need at least 205 MCQs/group (or 14 respondents) to have at least 90% power. Linear mixed models were used to compare scores between the 2 groups. Random effects for each of the 30 questions and 31 responders were used to account for the clustering. To assess whether respondents could differentiate AI-generated and human-generated MCQs, we used Chi-square test, and logistic regression with generalized estimating equations (GEE) to account for repeated measures from the same respondent. SAS software version 9.4 (Cary, NC, USA) and R version 4.4.2 (R Foundation for Statistical Computing, Vienna, Austria) was used for analysis.

### Survey Responders

The survey was distributed to 74 physicians who were rigorously vetted as experts in MV. All teach MV at academic institutions in the US. Their expertise is further validated by a specialized credentialing pathway: Each completed an intensive mechanical ventilation certification course, followed by a year-long advanced training program to qualify as preceptors in the field. This culminated in a 2.5-hour oral examination, where they earned highest evaluations—reflecting mastery of both theoretical knowledge and practical application in mechanical ventilation.

## Results

A total of 31 respondents (42%) completed the survey and evaluated the quality of the 30 MCQs. Their responses were analyzed to compare the quality of questions from both sources. The pre-specified non-inferiority margin was set at 15% of the maximum score (23), which corresponds to −3.45. Based on model estimates, the 95% one-sided upper confidence interval (CI) for the final score was calculated as (−1.15, ∞). Thus, non-inferiority criteria are confirmed indicating that AI-generated questions perform comparably to those created by human experts. Across all measures, there was no meaningful difference between AI-generated and human-expert MCQs. The mean values for final scores were nearly identical (AI vs. HE: −0.23 ± 0.56, p=0.67). Likewise, both groups performed similarly in terms of difficulty level, alignment with learning objectives, clarity of the question stem, correctness of the selected answer, and the plausibility of distractor options (Figure 1).

**Figure 1.**
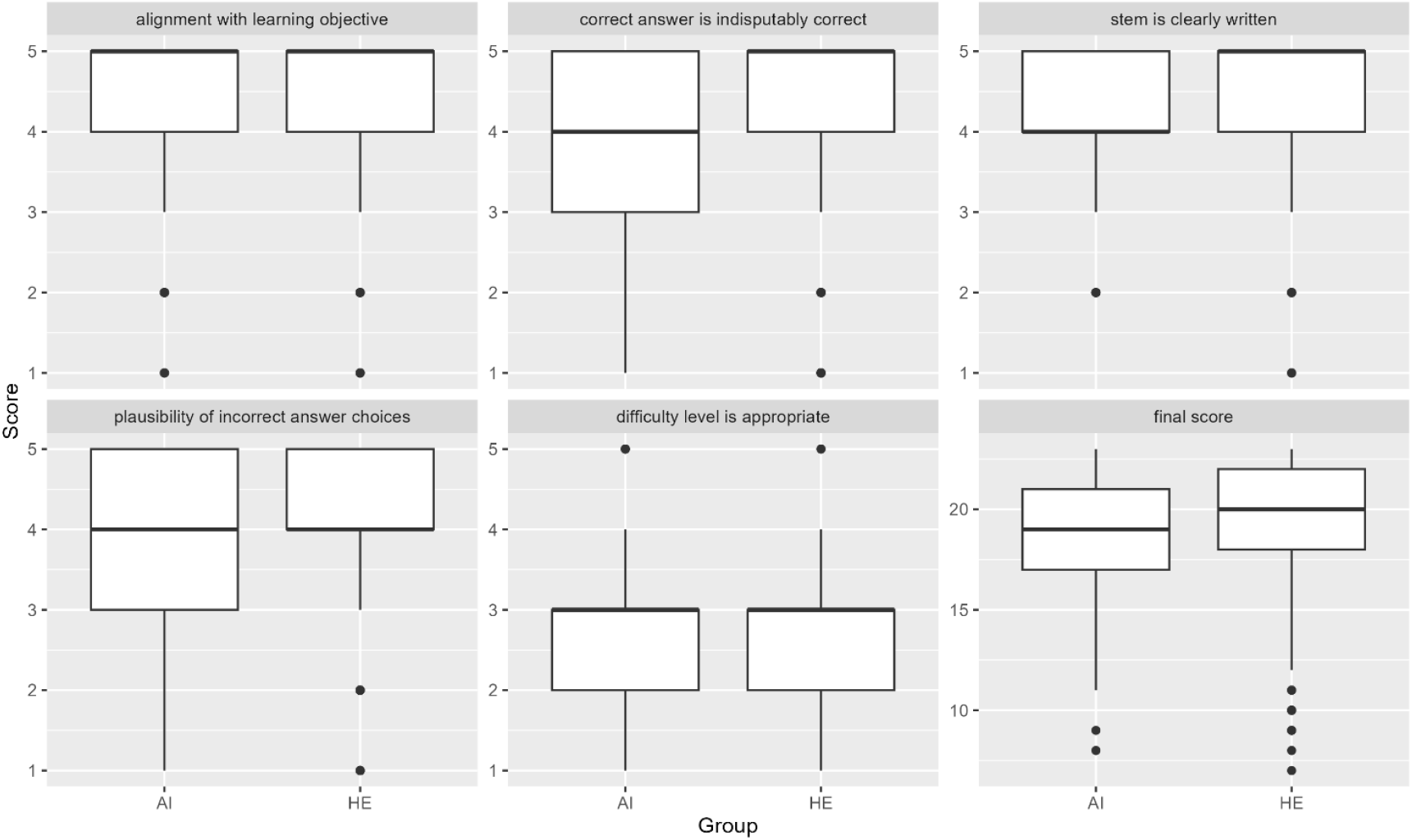
box plots showing survey responses by group. Distribution of scores of MCQ across the different domains of the survey grouped by the source of the MCQ, AI for aritifical intelligence, and HE for human expert

Finally, we looked at whether survey responders could differentiate between AI-generated and human-written questions. The respondents guessed 55.1% of the AI-generated MCQs correctly but also estimated 51.8% of the human-generated MCQs to be AI-generated (p=0.32, Chi-square test) (Table 2). The logistic regression analysis also did not show the guess to be a significant predictor of whether an MCQ is AI-generated (p = 0.32). Based on these results, we concluded that the survey responders were unable to reliably distinguish between AI-generated and human-written questions.

**Table 2.** Raters’ Ability to Identify Source of Multiple-Choice Question.

|  | GUESS -<br>ARTIFICIAL<br>INTELLIGENCE | GUESS -<br>HUMAN<br>EXPERT | TOTAL |
| --- | --- | --- | --- |
| **GROUP -**<br>**ARTIFICIAL**<br>**INTELLIGENCE** | 256 (55.1%) | 209 (45.0%) | 465 |
| **GROUP - HUMAN**<br>**EXPERT** | 241 (51.8%) | 224 (48.2%) | 465 |
| **TOTAL** | 497 | 433 | 930 |

## Discussion

Our study demonstrates that Chat-GPT, a widely available generative-AI LLM, produces high-quality MCQs for mechanical ventilation education comparable to those made by human educators and suitable for advanced fellowship training levels. The AI-generated MCQs performed comparably and were noninferior to human-developed questions across key assessment parameters, including difficulty level, alignment with learning objectives, and clarity of question-stem. In fact, the surveyed mechanical ventilation experts could not reliably distinguish AI vs human-generated questions.

HE-written MCQs were not significantly more consistent or accurate. Our findings, which demonstrate no significant advantage of HE-written questions over AI-generated questions, contrast with the findings of a prior study by Cheung et al. (2023). [8]

Advancements in LLM technology may account for the differences in results. However, variations in prompt design and delivery could also influence the final output. Recent research evaluating LLM performance against the American Academy of Orthopedic Surgeons (AAOS) osteoarthritis guidelines highlights the potential of prompt-engineering strategies to enhance model performance. [2] While not a direct comparison, the ChatGPT-o1 model used in our study demonstrates reasoning abilities that can be likened to executing a multi-step prompt.

Both possibilities need to be investigated further given the implications it could have on medical education. Our findings suggesting non-inferiority of AI-generated questions compared to those created by HE is particularly notable given the considerable time and resources required from human experts to create high-quality questions, particularly for a highly technical skill such as MV.

Our findings indicate that AI-generated questions can serve as a viable alternative to traditional question development, particularly in contexts where operational efficiency and scalability are critical. Mechanical ventilation is a highly technical subject area with limited accurate information available online; therefore, it is remarkable that the LLM-generated questions in our study were found to be non-inferior given that a specialized corpus of knowledge was not used in the creation of the AI-generated questions. While out of the scope of the data collected in our study, we believe that human expertise remains essential for quality assurance, especially in the creation of high-stakes assessments. Furthermore, LLM-generated content carries a risk of hallucination and misinformation. This is particularly concerning when creating MCQs in subject areas where unreliable information that influences the model’s training data is prevalent online, underscoring the need for human expert review of AI-generated questions.

Our study has several strengths. This is the largest study to date assessing AI-generated MCQs within the realm of medical education. The criteria used to assess the quality of the questions was robust as it was derived from consensus recommendations and guidelines from American Psychology Association (APA), National Council on Measurement in Education (NCME), and American Education Research Association (AERA). [14] Furthermore, our use of Chat-GPT to create our AI-generated questions is important because it demonstrates that MCQ generation can be done effectively using a widely available generative AI tool that does not require extensive specialized technical skills. Our meticulous non-inferiority margin selection a priori also reduced the probability of Type 1 error in our study. Additionally, the respondents of our survey were high-level experts in the subject area of mechanical ventilation, ensuring that the assessment of our MCQ’s was especially robust.

Our study has several weaknesses. First, out of a total of 74 possible respondents, 31 fully completed the survey, which is a response rate of 42%. Second, our AI-generated questions did not undergo further revision by human experts. AI-generated MCQs would most likely undergo further revision by human educators before official use in real-world educational settings, which would further enhance the quality of AI-generated MCQs relative to those solely created by humans. This suggests that our findings probably underestimate the potential quality of AI-generated questions that would ultimately be used in practice.

There are several promising areas for future exploration. In this study, we intentionally used a relatively simple prompt to reflect how an educator might use ChatGPT to generate questions online. However, since ChatGPT is not an open-source platform, future research could explore the use of open-source models. Unlike closed-source models, which operate on proprietary and confidential code with limited customization, open-source models provide users with the ability to view, modify, and share the underlying code. This openness enhances transparency, flexibility, and collaborative development. Additionally, these models can be run locally, offering benefits in terms of data privacy and control. However, they require greater technical expertise for installation, maintenance, and optimization.

One challenge with open-source language models is their variable performance quality. While some may not match the capabilities of proprietary models out of the box, fine-tuning can significantly enhance their effectiveness. [15] Additionally, generating questions based on new content may necessitate advanced techniques such as fine-tuning or retrieval-augmented generation (RAG), both of which require specialized knowledge. Our study provided valuable and novel insights into the potential real-world application of LLM’s for educators who do not possess specialized expertise in computer science or AI, a group that is probably representative of most medical educators.

Finally, while our study did not employ advanced prompt engineering, we recognize it as a growing field with the potential to enhance LLM performance. Prompt engineering focuses on refining input design to generate more accurate and reliable responses, with notable success in computer science, particularly in solving mathematical problems. Techniques like Chain of Thought (COT) and Tree of Thought (TOT) prompting techniques have shown improvements in computational tasks, suggesting that similar approaches could enhance LLM applications in other fields, such as medicine. [2] However, research into the impact of prompt engineering on medical applications is still in its early stages, with few studies evaluating how different prompt techniques influence LLM responses to medical queries.

## Conclusion

In conclusion, multiple choice questions generated by ChatGPT model o1 were comparable to that of human experts in the specific educational context of mechanical ventilation assessment. This supports the potential role of generative-AI in medical education assessment development.

## Data Availability

All data produced in the present study are available upon reasonable request to the authors

---

**Supplemental Table 1.** Survey Responses per Group.

Supplemental Table - Survey Responses per Group
| VARIABLE | ARTIFICIAL INTELLIGENCE | HUMAN EXPERT | EST DIFF (SE) | P-VALUE | 95% UPPER CI |
| --- | --- | --- | --- | --- | --- |
| OBJECTIVE ALIGNMENT (MEAN ± SD) | 4.34 ± 0.87 | 4.35 ± 0.94 | -0.002 (0.17) | 0.99 | (-0.28, ∞) |
| STEM CLARITY (MEAN ± SD) | 4.29 ± 0.85 | 4.32 ± 0.87 | -0.02 (0.12) | 0.85 | (-0.21, ∞) |
| CHOSEN ANSWER CORRECTNESS (MEAN ± SD) | 4.02 ± 1.17 | 4.22 ± 1.10 | -0.20 (0.24) | 0.40 | (-0.59, ∞) |
| INCORRECT ANSWERS PLAUSIBILITY (MEAN ± SD) | 3.88 ± 1.11 | 3.93 ± 1.16 | -0.05 (0.10) | 0.63 | (-0.22, ∞) |
| DIFFICULTY LEVEL (MEAN ± SD) | 2.68 ± 0.77 | 2.67 ± 0.81 | 0.01 (0.13) | 0.92 | (-0.21, ∞) |
| FINAL SCORE (MEAN ± SD) | 19.03 ± 2.94 | 19.26 ± 3.27 | -0.23 (0.56) | 0.67 | (-1.15, ∞) |

## Notes

### Competing Interest Statement

The authors have declared no competing interest.

### Funding Statement

This study did not receive any funding

